# Rethinking Measurement of Movement-Evoked Pain with Digital Technology

**DOI:** 10.1101/2025.09.14.25335734

**Authors:** Madelyn R. Frumkin, Jingwen Zhang, Ziqi Xu, Salim Yakdan, Braeden Benedict, Saad Javeed, Justin Zhang, Kathleen Botterbush, Burel R. Goodin, Chenyang Lu, Wilson Z. Ray, Jacob K. Greenberg

## Abstract

Movement-evoked pain (MEP) may be a useful metric for phenotyping musculoskeletal pain conditions. However, there is significant disagreement over operationalization, and no studies have assessed stability of MEP over time. Fitbit and Ecological Momentary Assessment (EMA) data were collected from adults with moderate-to-severe chronic pain schedule to receive lumbar/thoracolumbar fusion surgery (N=114). On average, participants provided 323 hours of Fitbit data and 74 EMA surveys (84% completion rate). To mimic task-based assessment of MEP using the 6-minute walk test, EMA pain ratings completed within 3 hours of walking at a speed ≥70spm for at least 6 minutes were extracted. Of the full sample, 91 individuals (80%) had any instances of pain ratings following 6-minute activity bouts (*Median*=6, *SD*=11). Post-activity pain scores exhibited good within-person consistency (ICC=.76). However, between-person differences in average pain accounted for >70% of the variance in post-activity pain. MEP change scores defined as the difference between post-activity and pre-activity pain scores had poor reliability (ICC = .08). MEP change scores were not associated with average pain or factors related to the uncontrolled nature of digital assessment (e.g., activity amount, time from activity to pain report). However, MEP change scores tended to be lower when the preceding pain rating was elevated (β = -7.96, 95% Credible Interval: -9.28, -6.66), suggesting ceiling effects. Small effects of time of day and prior activity were also observed, which could contaminate MEP assessed in the lab or clinic. Continued development of digital methodologies for assessing MEP is recommended.

**Perspective:** Existing movement-evoked pain assessments have limitations. Post-activity pain ratings capture overall disability and day-to-day fluctuations in pain, rather than the relationship between movement and pain. Pre-to-post activity change scores had poor reliability when assessed naturalistically and over time. Digital methodologies capture movement-evoked pain continuously across time, contexts, and real-world environments.

## 1. Introduction

There is growing emphasis on assessing musculoskeletal pain experienced with movement, or movement-evoked pain (MEP), as distinct from pain at rest (PAR)^9,10,12,21,22,58^. MEP tends to be greater than PAR^4,5^, and some studies find MEP is a stronger predictor of prospective outcomes, such as 12-month low back pain-related disability and early recovery after spine surgery^7,8^.

However, findings are highly mixed. For example, in knee arthroplasty, some studies suggest MEP and PAR are distinct^9,10^, while others describe MEP and PAR as substantially overlapping^11^.

Mixed findings may be due to heterogeneity in the methods used to assess MEP. MEP can be assessed retrospectively by asking individuals to rate their pain with activity over a recent time period (e.g., the past 24 hours or the past week). However, retrospective recall of pain is known to be influenced by factors including the individual’s most intense and most recent pain experiences^12–14^. It is therefore suggested that MEP be assessed in response to tasks likely to evoke pain (e.g., six-minute walk test, repeated chair rise)^3,4^. In some cases, a sequential battery of tasks is used to characterize aggregate MEP (e.g., ^15–17^).

A key discrepancy in task-based assessment of MEP is whether MEP should be operationalized as absolute post-task pain versus the degree to which pain increases following activity^2^.

Numerous studies have used absolute post-activity pain ratings to quantify MEP, such that individuals who report higher post-activity pain (e.g., pain after walking) are considered to have greater MEP. Absolute post-activity pain tends to be highly correlated with PAR and overall disability^8,11,16,18^. Prior Ecological Momentary Assessment (EMA) research suggests that pain is highly variable within individuals over time, due to factors including negative cognitive and affective states^19,20^, sleep^21^, and time of day^22,23^. Because task-based assessments of MEP are typically performed on only one occasion, it is unclear if post-activity pain ratings are similarly variable, whether due to pre-activity pain or independent influences of these factors on post-activity pain ratings.

Alternatively, MEP can be isolated from PAR by calculating the difference between pre- and post-activity pain^2^. Such MEP change scores characterize change in pain following one or multiple tasks and are intended to index the degree to which pain increases with activities expected to evoke pain for a given patient population. Similarly to absolute post-activity pain, it is possible that the degree to which pain increases following activity is variable over time due to factors including pre-activity pain level, time of day, and how much activity the individual has engaged in prior to MEP assessment^24–26^. For example, difference scores may suffer from ceiling effects if the individual experiences high resting pain prior to engaging in activity intended to evoke pain.

The goal of the current study was to leverage digital technology to assess stability of MEP assessments over time and under naturalistic conditions. To mimic task-based assessment via the six-minute walk test, EMA pain ratings preceded by physical activity bouts of at least six-minutes were extracted. Reliability of both absolute and change score metrics of MEP were examined. The degree to which MEP metrics were influenced by average pain, pre-activity pain, factors related to the uncontrolled nature of digital assessment (e.g., activity amount, time from activity to pain report), and factors that could contaminate MEP assessed in the lab or clinic (e.g., time of day, prior activity) was further assessed in univariate and multivariate models. Finally, the risk of ceiling effects was assessed by naturalistically examining PAR following periods of no or low physical activity.

## 2. Materials and methods

### 2.1. Participants

This is a secondary analysis of data collected to evaluate the feasibility and utility of preoperative mobile health (mHealth) assessment for improving prediction of lumbar spine surgery outcomes. The methods and primary outcomes have been reported elsewhere^7,27^. Briefly, inclusion criteria included English-speaking adults aged 21 to 85 years old who owned a smartphone, had at least 1 week to complete assessments prior to surgery, and reported a numeric rating scale pain score of at least 3 out of 10 during the previous week. Patients who were undergoing surgery for infection, malignancy, or trauma, those undergoing isolated thoracic fusion, and those undergoing another major surgery within 3 months of data collection were excluded. The study was approved by the institutional review board at Washington University School of Medicine (IRB# 202012139), and all patients provided informed consent. To be included in the current study, participants were also required to have both EMA and Fitbit data available preoperatively. At present, neither patients nor the public have been involved in the study’s conceptualization, design and conduct (e.g., choice of outcome measures, recruitment), or dissemination of the study results.

### 2.2. Procedure

Participants were recruited over the phone following a recent appointment with a neurosurgeon or orthopedic spine surgeon. A research coordinator contacted the patient, explained the study, and assessed interest in further participation. If participants indicated interest in the study, then the research coordinator verbally reviewed the purpose and procedures of the study, the voluntary nature of participation, access to protected health information, and study compensation. If patients indicated they would like to participate, they then provided informed consent and were given instructions to download the LifeData application (LifeData LLC) to complete EMA on their personal smartphone. Participants specified a 12-hour period in which they preferred to receive surveys every 3 hours (i.e., 9am to 9pm). Participants also completed self-report questionnaires via REDCap.

After enrolling in the study, participants were mailed a Fitbit Inspire 2 with instructions to wear the tracker as much as possible but at least during the 12-hour EMA period. Participants received 5 EMAs daily for approximately 3 weeks, or until their surgery. Some individuals experienced delays in surgery date due to COVID-19, illness, or other factors. For consistency across participants, analyses focus on data collected within 45 days of surgery. Participants could choose the time at which their surveys started. EMA surveys were administered every 3 hours^1^.

The EMA schedule was fixed, such that participants received surveys at the same times for the duration of the study (e.g., 9am, 12pm, 3pm, 6pm, 9pm). EMAs were not triggered by activity or other variables. Participants had 30 minutes to respond to each survey and were sent 2 reminders at 15-minute increments. Participants were paid $1 per completed EMA survey (up to $105), and $20 for using the Fitbit for any duration (see ^27^ for further details).

### 2.3. Measures

#### 2.3.1. Ambulatory movement assessment

Activity data were collected via the Fitbit Inspire 2. Fitbit provides step count and heart rate data extracted every minute. Validity or reliability of Fitbit-based measurements have been assessed in at least 144 studies, and results of a systematic review suggest that Fitbits accurately measure steps^28^.

#### 2.3.2. Momentary pain

At each scheduled EMA assessment, participants rated pain intensity (“Right now, how intense is your overall pain?”) on a scale from 0 (none) to 100 (worst possible). Participants were instructed to respond based on how they were feeling right before they received the notification. Participants were not given instructions as to whether to reference pain with movement or rest.

#### 2.3.3. Patient-reported outcome measures

Participants completed computer-adaptive Patient-Reported Outcomes Measurement Information System (PROMIS) pain intensity, pain interference, and physical function measures upon entering the study^29–31^. PROMIS scores are reported as t-scores, where 50 corresponds to population average, and 10 is the standard deviation. Higher scores on PROMIS pain intensity and interference and lower scores on physical function suggest impairment^29^.

### 2.4. Data preparation

#### 2.4.1. Data cleaning

Fitbit records step count as zero even if the device is not worn. Therefore, step count observations were removed if heart rate data were not available.

#### 2.4.2. Movement-evoked pain

A schematic overview of ecological MEP assessment is provided in Figure 1A. Pain ratings were marked as indexing MEP if the individual engaged in a physical activity (PA) bout of at least 6 minutes in the 3 hours prior to the pain rating. This time period was chosen to mimic the six-minute walk test (6MWT), which is commonly used to measure MEP in laboratory and clinical settings^3^. The 3-hour window was chosen because pain was measured every 3 hours via EMA. Based on prior literature, PA was defined based on a threshold of ≥70 steps per minute (spm)^32^.

**Figure 1.**
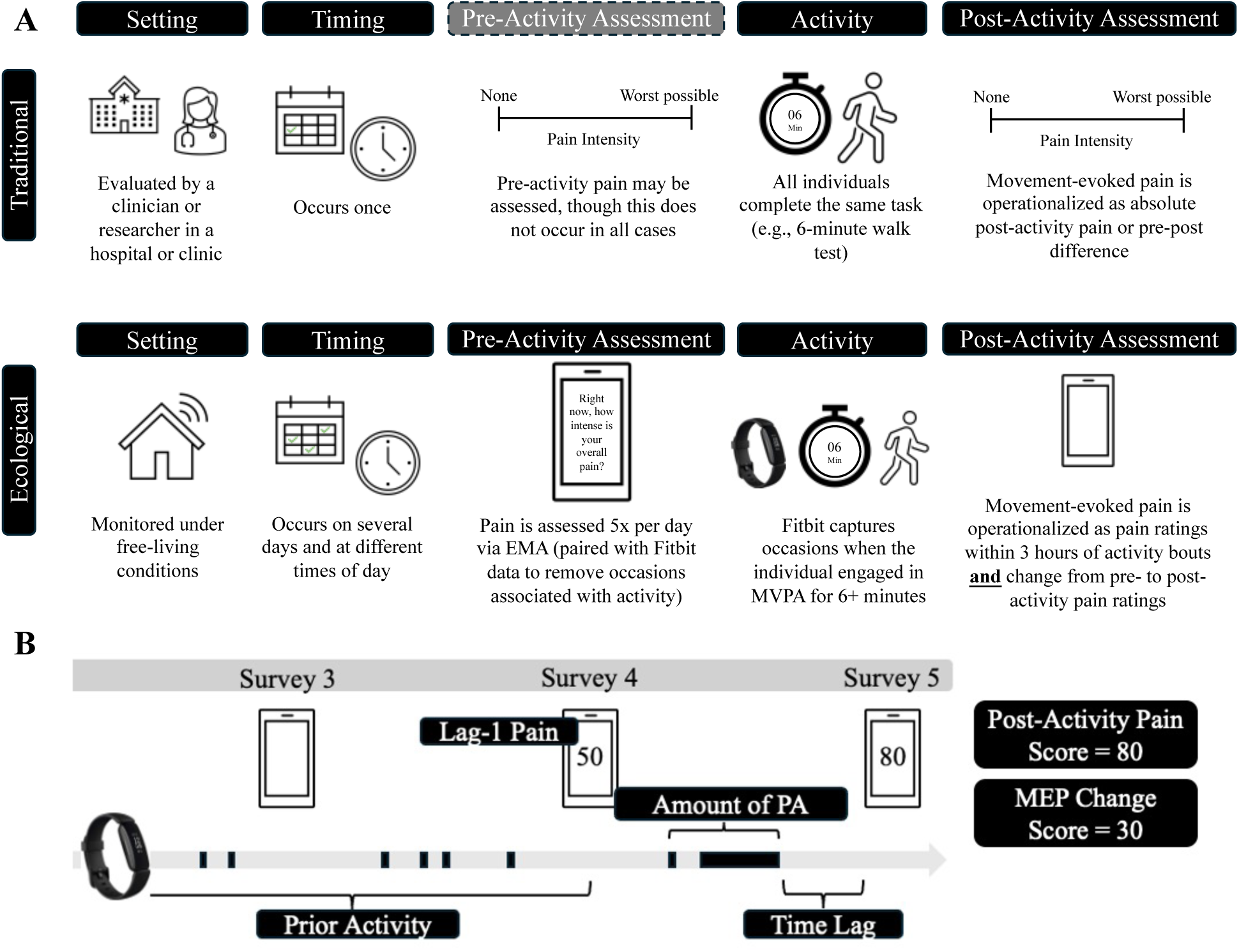
Schematic overview of traditional versus ecological movement-evoked pain assessment (**A**) and time-varying predictors of ecological MEP observations (**B**). EMA = ecological momentary assessment; PA = physical activity.

Several step count thresholds were considered to confirm that heart rate was sufficiently elevated to suggest at least light-to-moderate PA (see 2.5.1).

MEP is defined in the literature as either absolute post-activity pain ratings or the change in pain from pre- to post-activity. Both operationalizations are included in this analysis. In the first set of models, the time-varying outcome is EMA pain ratings following bouts of PA (henceforth referred to as “post-activity pain”). In the second set of models, the time-varying outcome is the difference between EMA pain ratings following bouts of PA and lag-1 EMA pain ratings made approximately 3 hours earlier by the same individual (henceforth referred to as “MEP change score”). MEP change score was calculated only if the lag-1 EMA pain rating was not associated with a bout of PA, so that lag-1 pain more closely mimicked PAR.

#### 2.4.3. Predictors of MEP

Raw Fitbit data and EMA pain ratings were used to generate secondary features for predicting post-activity pain. Predictors are described below and in Figure 1B.

##### Average pain

Average pain was calculated for each participant as the average of that individual’s momentary pain responses, regardless of whether Fitbit data were available. The number of EMA responses available for calculating average pain varied based on EMA compliance and number of days enrolled prior to surgery (see Table 1). However, the minimum was 9 observations, which is likely sufficient for reliable estimation of the person-level mean^33,34^. For each participant, an average across all EMA responses was calculated. Person-level means were then grand-mean centered and scaled, such that the coefficient reflects the expected change in average MEP for an individual whose average pain is 1 SD above the sample mean.

**Table 1.**
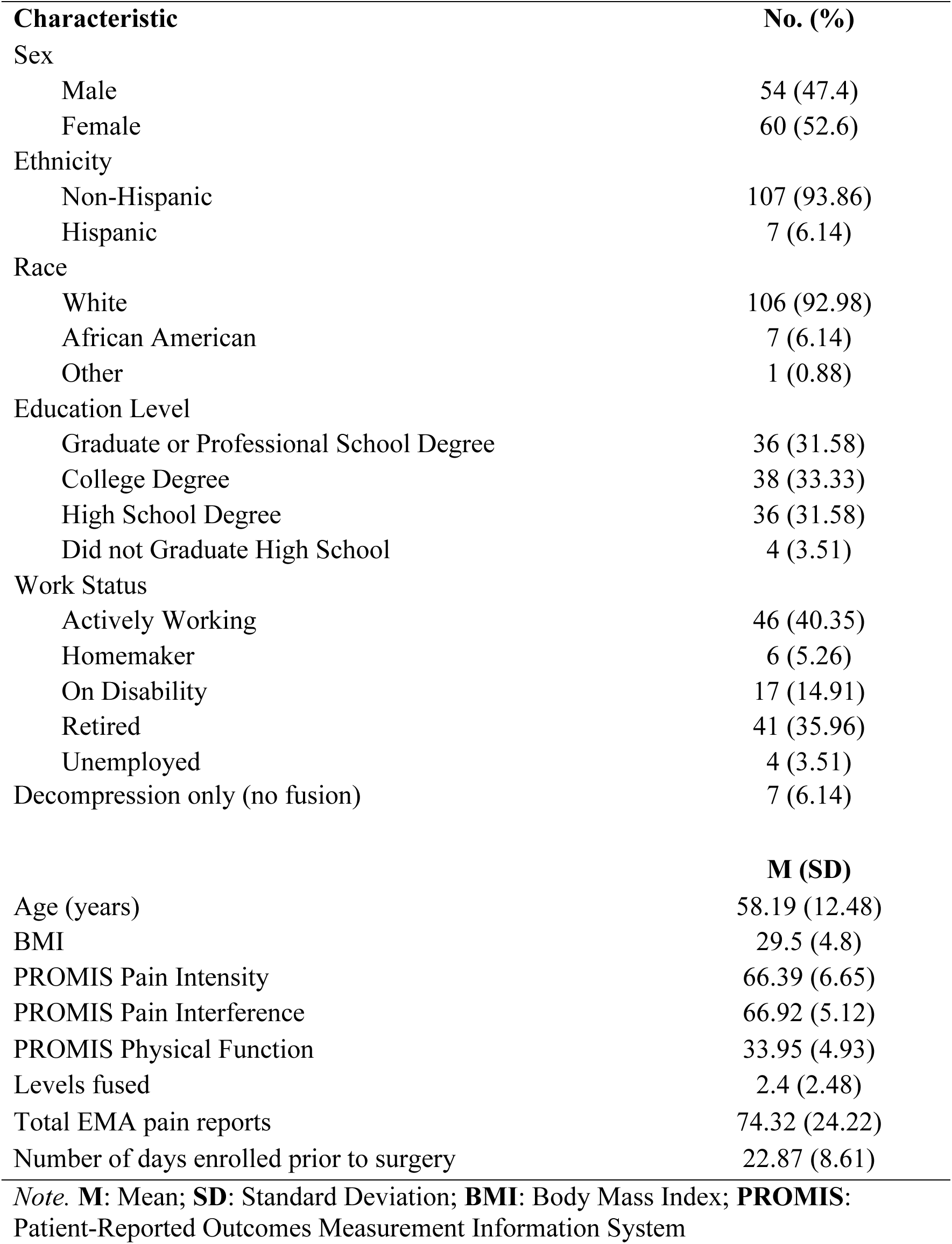
Sample descriptives (N = 114)

| **Characteristic** | **No. (%)** |
| --- | --- |
| Sex |  |
| Male | 54 (47.4) |
| Female | 60 (52.6) |
| Ethnicity |  |
| Non-Hispanic | 107 (93.86) |
| Hispanic | 7 (6.14) |
| Race |  |
| White | 106 (92.98) |
| African American | 7 (6.14) |
| Other | 1 (0.88) |
| Education Level |  |
| Graduate or Professional School Degree | 36 (31.58) |
| College Degree | 38 (33.33) |
| High School Degree | 36 (31.58) |
| Did not Graduate High School | 4 (3.51) |
| Work Status |  |
| Actively Working | 46 (40.35) |
| Homemaker | 6 (5.26) |
| On Disability | 17 (14.91) |
| Retired | 41 (35.96) |
| Unemployed | 4 (3.51) |
| Decompression only (no fusion) | 7 (6.14) |
|  | **M (SD)** |
| Age (years) | 58.19 (12.48) |
| BMI | 29.5 (4.8) |
| PROMIS Pain Intensity | 66.39 (6.65) |
| PROMIS Pain Interference | 66.92 (5.12) |
| PROMIS Physical Function | 33.95 (4.93) |
| Levels fused | 2.4 (2.48) |
| Total EMA pain reports | 74.32 (24.22) |
| Number of days enrolled prior to surgery | 22.87 (8.61) |

*Note.* **M:** Mean; **SD:** Standard Deviation; **BMI:** Body Mass Index; **PROMIS:** Patient-Reported Outcomes Measurement Information System

##### Time lag

In this ecological setting, pain ratings were not requested immediately after activity bouts. Time (in hours) from end of the PA bout to the EMA pain rating was calculated. The maximum possible time lag was 3 hours. If more than one activity bout occurred in the 3-hour window, time lag refers to time since the end of the most recent bout. The coefficient reflects the expected change in MEP per 1-hour increase in time lag.

##### Survey

Given that pain can fluctuate with circadian rhythms^23,35^, survey number was included as a predictor of MEP. Surveys 1-5 were delivered at the same time each day for a given participant. Both linear and quadratic (survey^2^) effects were examined after recoding survey so that 0 referenced the mid-day (e.g., 3pm) survey.

##### Lag-1 pain

EMA pain ratings made approximately 3 hours before a post-activity pain observation by the same individual were included as a time-varying predictor. Sequential MEP observations were removed so that lag-1 pain more closely mimicked PAR. Remaining lag-1 pain observations were not associated with 6+ minute activity bouts, based on the step count threshold. However, it was not required that participants be completely sedentary prior to the lag-1 pain observation, as this would increase the amount of missing data. The impact of this decision was evaluated in subsequent analyses (see 2.5.4.). Lag-1 pain was centered and scaled within persons, such that the coefficient reflects the expected change in MEP when lag-1 pain is 1 SD above average for the individual.

##### Amount of PA

Individuals may have engaged in PA for differing amounts of time across bouts, and multiple bouts may have occurred within the 3-hour window. Total number of minutes above the PA threshold in the 3 hours prior to a post-activity pain rating was calculated. Amount of PA was centered and scaled within persons, such that the coefficient reflects the expected change in MEP when amount of PA within the 3-hour window is 1 SD above average for the individual.

##### Prior activity

Because MEP may be additive, amount of daily activity prior to the MEP occasion was included as a predictor. To avoid conflating prior activity with amount of PA, this variable was operationalized as cumulative step count up to 3 hours before the post-activity pain rating (e.g., if the post-activity pain rating was recorded at 9pm, cumulative step count up to 6pm indicated prior activity). To account for times when participants may not have worn the Fitbit, cumulative step count was divided by cumulative wear time. Prior activity was centered and scaled within persons, such that the coefficient reflects expected change in MEP when prior activity is 1 SD above average for the individual.

### 2.5. Statistical Analysis

#### 2.5.1. Defining physical activity bouts

Various thresholds have been used to define sedentary vs. active time using Fitbits and other wearable devices^36,37^. Fitbit uses a proprietary algorithm to categorize 60-second sampling intervals as sedentary, light, moderate, or vigorous activity. However, active time is over-estimated compared to research-grade devices^38,39^. Physical activity bouts were therefore quantified based on step count thresholds. Guidelines suggest that moderate intensity walking involves ≥100 steps per minute (spm), including among older adults^40^. However, such a cadence may be uncommon in individuals with physical disability, and lower walking speeds (e.g., 70 steps per minute) may be sufficient for expanding 3 metabolic equivalents (METs), a common threshold for moderate-intensity PA^32,37,40–42^.

Given a lack of prior research defining physical activity intensity among adults with moderate-to-severe pain interference and disability, thresholds of 50, 60, 70, 80, 90, and 100 spm were considered. For each threshold, all 6-minute windows when average spm was at or above the threshold were identified. This time period was chosen to mimic the 6MWT. Simultaneous heart rate data were then used to verify presence of moderate-intensity PA. Minute-level heart rate data was divided by the individual’s maximum heart rate (calculated as 208 - 0.7*Age) and averaged within each 6-minute window, yielding average percent of maximum heart rate for the activity bout^43^. The proportion of activity bouts where the individual’s heart rate was elevated to at least 50% and 64% of their maximum heart rate are reported. These reflect typical benchmarks for moderate-intensity physical activity^44^.

#### 2.5.2. Describing reliability of MEP

The Intraclass Correlation Coefficient (ICC) indexes the proportion of variance that is due to between-person differences. Person-specific standard deviation of each MEP metric (absolute post-activity pain and MEP change scores) are also reported.

#### 2.5.3. Examining multilevel predictors of absolute post-activity pain

Mixed-effects models were fit using the lmerTest package in R version 4.4.1^45^. All models include a random intercept, allowing for individual differences in average MEP. There were two time-varying outcomes: post-activity pain and MEP change score. For each outcome, univariate models were fit to understand the impact of potential predictors on both MEP operationalizations. Marginal R^2^ describe the proportion of variance accounted for by predictors of interest (i.e., fixed effects).

Univariate models use complete case analysis. Most predictors had no missing data, because they were process variables (e.g., time of day), aggregate variables (e.g., person-level average pain), or necessary for inclusion in the data set of MEP observations (e.g., Fitbit data). The exception is lagged pain, which was sometimes missing at the prior assessment (e.g., approximately 3 hours prior) and always missing at the first survey of the day. Missing observations were not imputed due to problems with parameter estimation when data are not missing at random and a high proportion of missing data are present^46^. Instead, multivariate models were fit using Bayesian multilevel dynamic structural equation modeling in Mplus. Through Kalman filtering, all available observations are included in the analyses, even if predictors are missing. Uncertainty due to missing values is incorporated into model estimates via posterior distributions^47,48^.

#### 2.5.4. Describing variability in resting pain

In the main analysis, PAR is defined as any lag-1 EMA pain observation that was not marked as indexing MEP (i.e., not preceded by an activity bout lasting 6+ minutes within 3 hours of the pain rating). Individuals were not required to be completely sedentary, as this would increase the amount of missing data, and MEP protocols vary in how PAR is assessed (e.g., amount of resting time). Instead, variability in PAR as a function of pre-EMA activity was empirically examined.

First, distributions in PAR and pre-EMA activity are examined across all EMA pain observation not marked as indexing MEP. This analysis was then restricted to lag-1 EMA pain observations included in the main analysis (i.e., those that preceded an activity bout and subsequent MEP observation). The proportion of observations where resting pain was reported as ≥ 80 on a 0-100 scale indicating severe pain^49^ is reported.

## 3. Results

Participants were 114 adults with moderate-to-severe pain interference and disability scheduled to receive lumbar spine surgery (see Table 1). On average, participants provided 323 hours (*SD* = 172) of Fitbit data over 18 days (*SD* = 8). When considering days when participants wore the Fitbit for at least 12 hours during waking hours (e.g., excluding 12am-6am), average daily step count was 6453 (*SD* = 3596, *Min* = 258, *Max* = 15788). Participants completed 84% of EMA prompts on average (*SD* = 15%), yielding an average of 74 completed prompts per person (*SD* = 24).

### 3.1. Defining physical activity bouts

Maximum steps per minute ranged from 50 to 158, with 11% of participants never exceeding 100 steps/minute (see Figure 2A). As shown in Figure 2B, average heart rate was elevated at the 50spm threshold (*M* = 102.55, *SD* = 9.97), compared to 6-minute windows when participants were unmoving (*M* = 74.48, *SD* = 11.27) or averaged <10spm (*M* = 76.74, *SD* = 11.29). Average heart rate continued to increase at higher step count thresholds. However, as the threshold increased, fewer participants had any qualifying 6-minute activity bouts (see Figure S1). Only 54 participants (47%) had any 6-minute windows with ≥100spm. The 70spm threshold was therefore retained. Results for alternative thresholds are presented in the supplementary materials.

**Figure 2.**
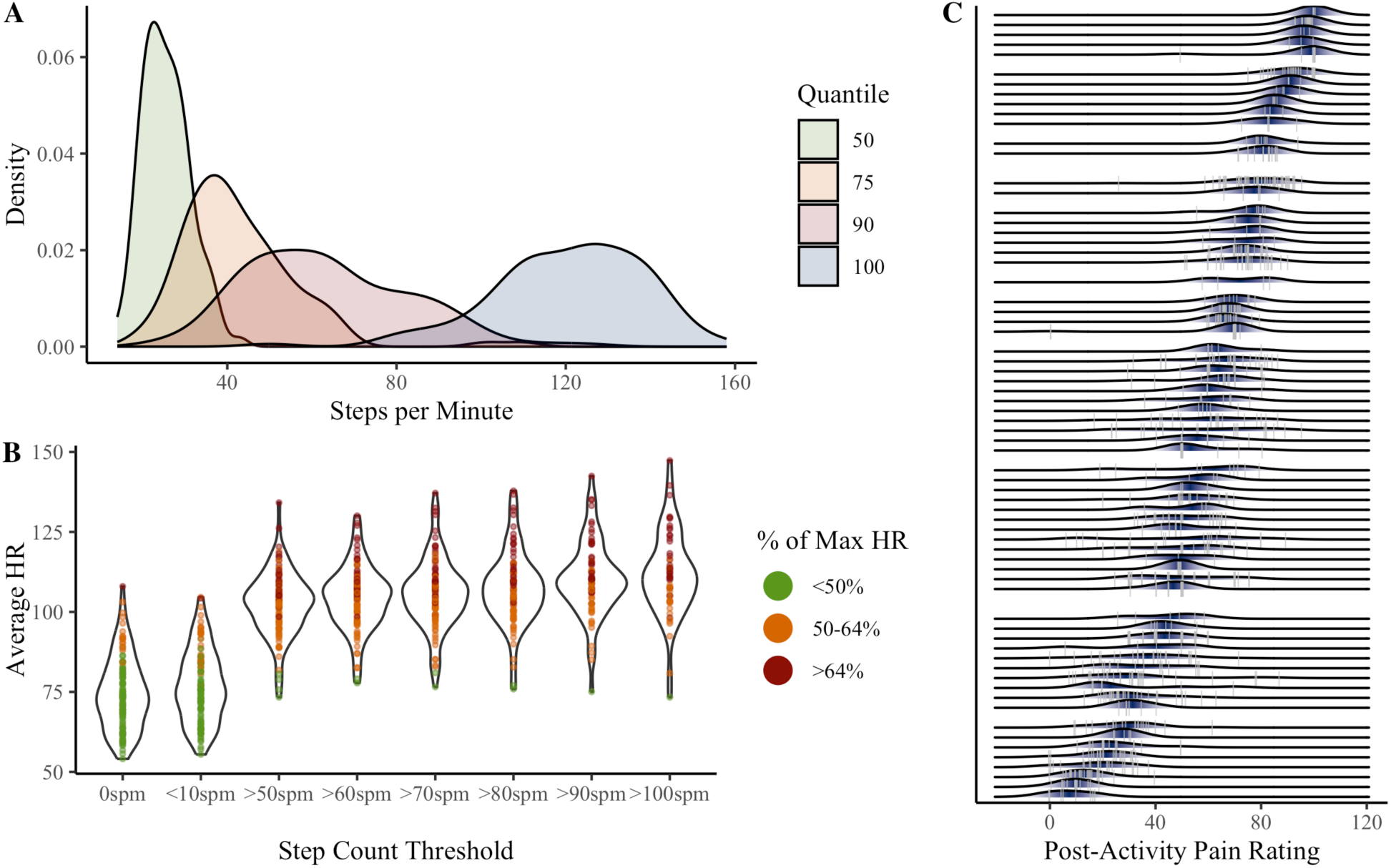
Variability in step quantiles (**A**), heart rate at step count thresholds (**B**), and movement-evoked pain ratings (**C**). Panel **A** shows between-person variability in step counts associated with the 50^th^, 75^th^, 90^th^, and 100^th^ quantiles during non-sedentary minutes (step count ≥ 10 steps per minute). 50^th^ quantile refers to the individual’s average (non-sedentary) step count; 100^th^ quantile refers to the individual’s maximum steps per minute. Panel **B** shows person-level average heart rate associated with various step count thresholds (spm = steps per minute). Dots indicate person-level average heart rate for each threshold, across all available 6-minute activity bouts. For each bout, percent of maximum heart rate was calculated as observed average heart rate divided by the individual’s maximum heart rate (208 - 0.7*Age). Panel **C** shows person-level variability in post-activity pain recorded within 3 hours of an activity bout (defined as at least 6 consecutive minutes walking at a speed ≥ 70 steps per minute). Grey vertical lines are observed post-activity pain ratings. Shaded areas are person-level probabilities (darker = higher probability).

### 3.2. Defining MEP

Of the full sample, 91 participants (80%) had any pain ratings following a 6-minute activity bout based on the 70spm threshold (T = 1022). 46 observations (4.5%) were removed because percent of maximum heart rate was <50%. Among individuals with any post-activity pain ratings, the median number of observations was 6 (*SD* = 11), corresponding to an average of 14% (*SD* = 13%) of the individual’s EMA pain ratings.

To define MEP, sequential observations (t = 270, 27.7%) were removed so that lag-1 pain more closely mimicked PAR. The remaining 715 post-activity pain observations across 90 individuals were used for the first operationalization of MEP (absolute post-activity pain). Calculating MEP change scores requires a lagged pain observation. 496 MEP change scores were thus retained across 85 individuals after removing observations that occurred at the first survey of the day when lag-1 pain was not available (t = 99, 45% of missing lag-1 pain occasions) or later in the day when lag-1 EMA was not completed (t = 120).

### 3.3. Reliability of MEP

Variability in post-activity pain is visualized in Figure 2C. The average within-person standard deviation of post-activity pain was 10.14 (*SD* = 6.64). The ICC of .76 suggests approximately 24% of the variance in post-activity pain occurred within individuals.

The average within-person standard deviation of MEP change score was 11.58 (*SD* = 7.78). The ICC of .08 suggests that approximately 92% of the variability in MEP change score occurred within individuals. Variability and ICC estimates were similar across step count thresholds (Table S1).

### 3.4. Univariate predictors of absolute post-activity pain

Results of the univariate mixed-effects models predicting post-activity pain are presented in Figure 3. Individuals with greater pain on average tended to report higher post-activity pain (β = 22.46, *p* < .001). Average pain accounted for 71.3% of the variance in post-activity pain. When examined over time, post-activity pain tended to be higher when lag-1 pain was above average for the individual (Fig 3B). These effects were consistent across step count thresholds (Table S2).

**Figure 3.**
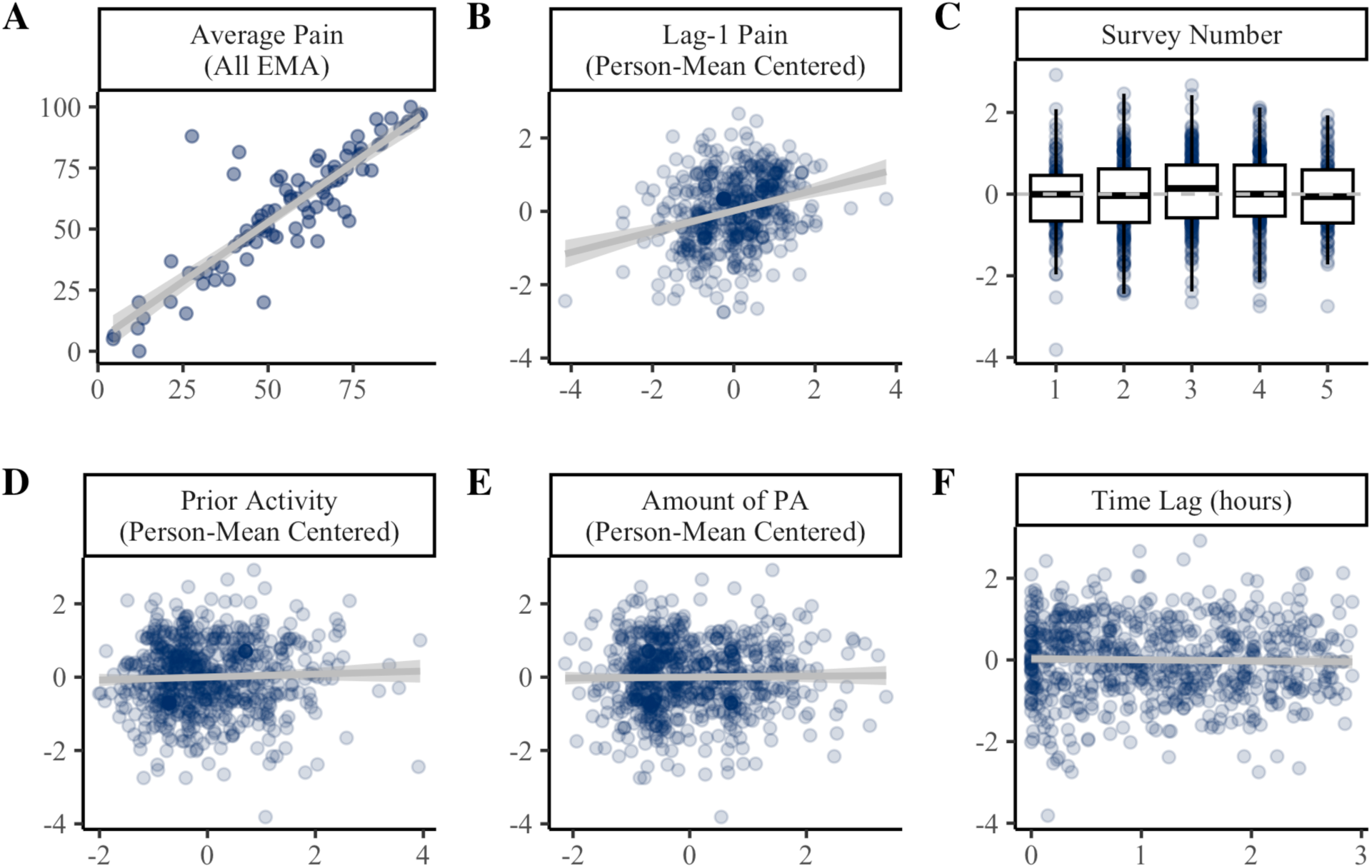
Predictors of post-activity pain ratings. Panel **A** shows the between-persons association of average pain across all EMAs (x-axis) with average post-activity pain (y-axis). Panels **B-F** show the effect of time-varying covariates (x-axis) on within-person variability in post-activity pain ratings (y-axis). In Panels B-F, post-activity pain is person mean centered such that 0 = average for the individual. PA = physical activity. Coefficients and standard errors are available in Table S2.

At the 70spm threshold, post-activity pain ratings were not significantly influenced by time of day (*p* = .067; Fig 3C). Notably, there was a quadratic effect of time at most other thresholds examined (Table S2), such that post-activity pain appeared highest at the mid-afternoon survey and lowest in the evening, though this accounted for <1% of variance in post-activity pain ratings. Post-activity pain ratings were not influenced by amount of activity prior in the day (Fig 3D), or the amount of activity within the 3-hour window (Fig 3E). EMA pain ratings occurred 1.2 hours after activity bouts on average (SD = .87 hours). Post-activity pain ratings were not associated with the time lag from activity to EMA (Fig 3F).

### 3.5. Multivariate predictors of absolute post-activity pain

Results of the multivariate model are presented in Table 2. Average pain continued to be the strongest predictor of post-activity pain. Consistent with univariate models, lag-1 pain also accounted for unique within-person variance in post-activity pain ratings over time. These findings were consistent across step count thresholds (Table S3). Time of day and cumulative activity prior to the lag-1 pain observation were also small but significant predictors based on Bayesian one-tailed p-values. However, the 95% credible intervals contained zero, suggesting effects are not reliably different from zero. Consistent with univariate models, there was a quadratic effect of time at other step count thresholds examined (Table S3). The multivariate model accounted for 97.1% of the between-person variance and 6.5% of the within-person variance in post-activity pain.

**Table 2.** Multivariate predictors of post-activity pain (N = 90, T = 715)

| Predictor | Estimate | Posterior SD | P | 95% CI |
| --- | --- | --- | --- | --- |
| **[Intercept]** | **59.971** | **0.892** | **<.001** | **[58.241, 61.773]** |
| **Average Pain** | **22.307** | **0.762** | **<.001** | **[20.753, 23.749]** |
| **Lag-1 Pain** | **3.310** | **0.645** | **<.001** | **[2.035, 4.562]** |
| Survey<sup>2</sup> | -0.632 | 0.331 | .030 | [-1.277, 0.023] |
| Amount of PA | -0.563 | 0.567 | .160 | [-1.676, 0.557] |
| Prior Activity | 1.120 | 0.576 | .026 | [-0.011, 2.248] |
| Time Lag | -0.424 | 0.623 | .247 | [-1.640, 0.800] |
| R<sup>2</sup> Between | 0.971 | 0.016 |  | [0.929, 0.993] |
| R<sup>2</sup> Within | 0.065 | 0.021 |  | [0.030, 0.113] |

**Note.** SD = Standard Deviation; P = Bayesian one-tailed p-value based on the posterior distribution; CI = Credible Interval; PA = physical activity

### 3.6. Univariate predictors of MEP change scores

Results of the univariate mixed-effects models predicting MEP change scores are presented in Figure 4. Average pain was uncorrelated with average MEP change score. When examined over time, MEP change score tended to be lower when lag-1 pain was elevated (β = -8.17, *p* < .001). Lag-1 pain accounted for 22% of the variance in MEP change score and was a consistent predictor across step count thresholds (Table S4).

**Figure 4.**
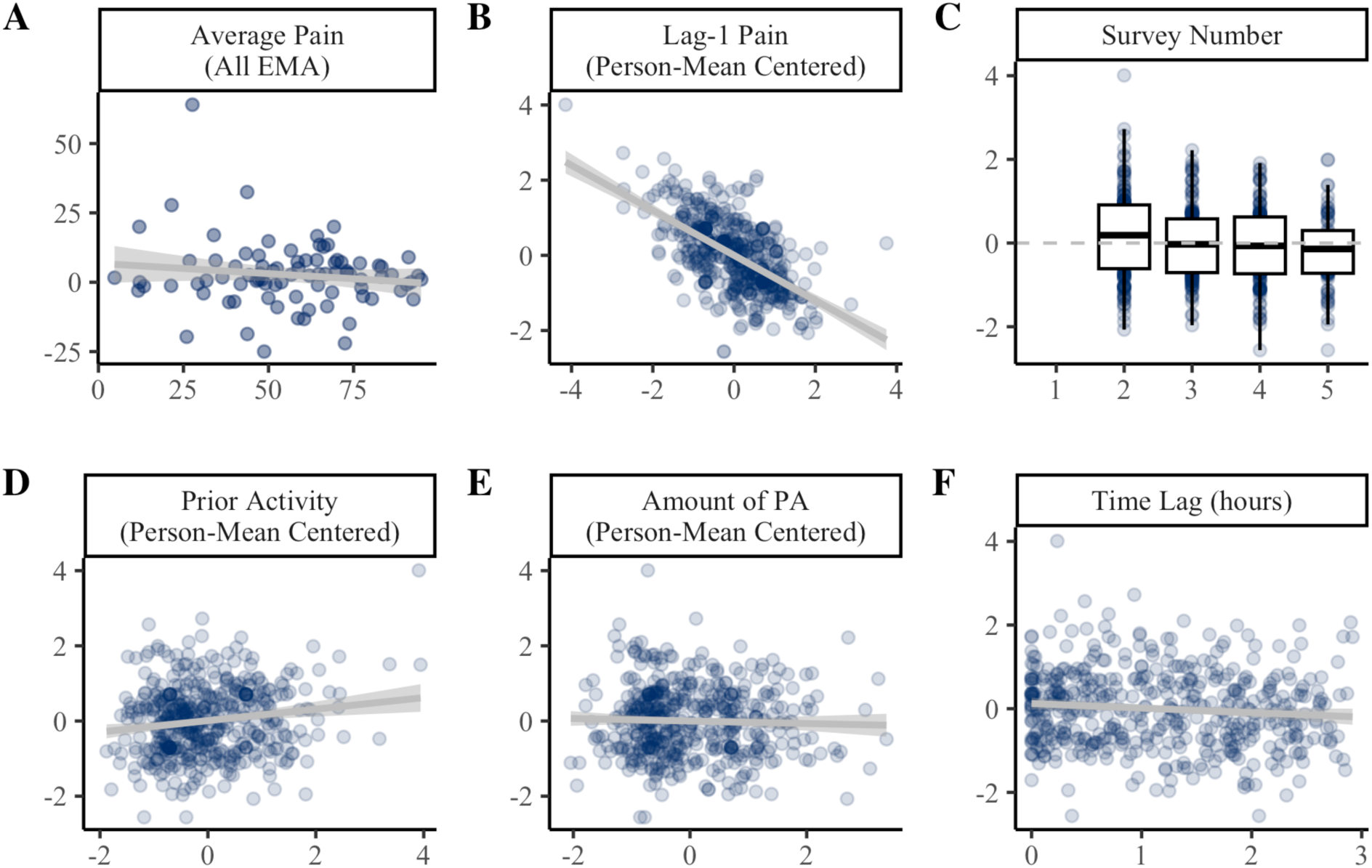
Predictors of MEP change scores. Panel **A** shows the between-persons association of average pain across all EMAs (x-axis) with average MEP change score (y-axis). Panels **B-F** show the effect of time-varying covariates (x-axis) on within-person variability in MEP change scores (y-axis). In Panels B-F, MEP change score is person mean centered such that 0 = average for the individual. PA = physical activity. Coefficients and standard errors are available in Table S4.

At the 70spm threshold, there was a linear effect of time, such that MEP change score tended to decrease across surveys 2 (e.g., 12pm) to 5 (e.g., 9pm; Fig 4C). MEP change scores also tended to be elevated when individuals had been more active so far that day (β = 2.05, *p* = .001; Fig 4D). Notably, time of day and prior activity were not consistent predictors across higher step count thresholds (Table S4).

MEP change scores were not influenced by the amount of PA within the 3-hour window (Fig 4E) or time lag from activity to EMA (Fig 4F). At lower step count thresholds (e.g., 50-60spm), MEP change scores tended to be lower when collected further from the time of activity.

### 3.7. Multivariate predictors of MEP change scores

Results of the multivariate model are presented in Table 3. Lag-1 pain accounted for unique variance in MEP change scores at all step count threshold (Table S5). Cumulative activity prior to the lag-1 pain observation was a small but significant predictor based on Bayesian one-tailed p-value. However, the 95% credible interval contained zero, suggesting effects are not reliably different from zero. Prior activity was a predictor of increased MEP change scores at the 60spm threshold (Table S5). In the multivariate models, increased time from activity bout predicted lower MEP change score only at the 50spm threshold. The multivariate model accounted for 2.1% of the between-person variance and 25.9% of the within-person variance in MEP change scores.

**Table 3.**
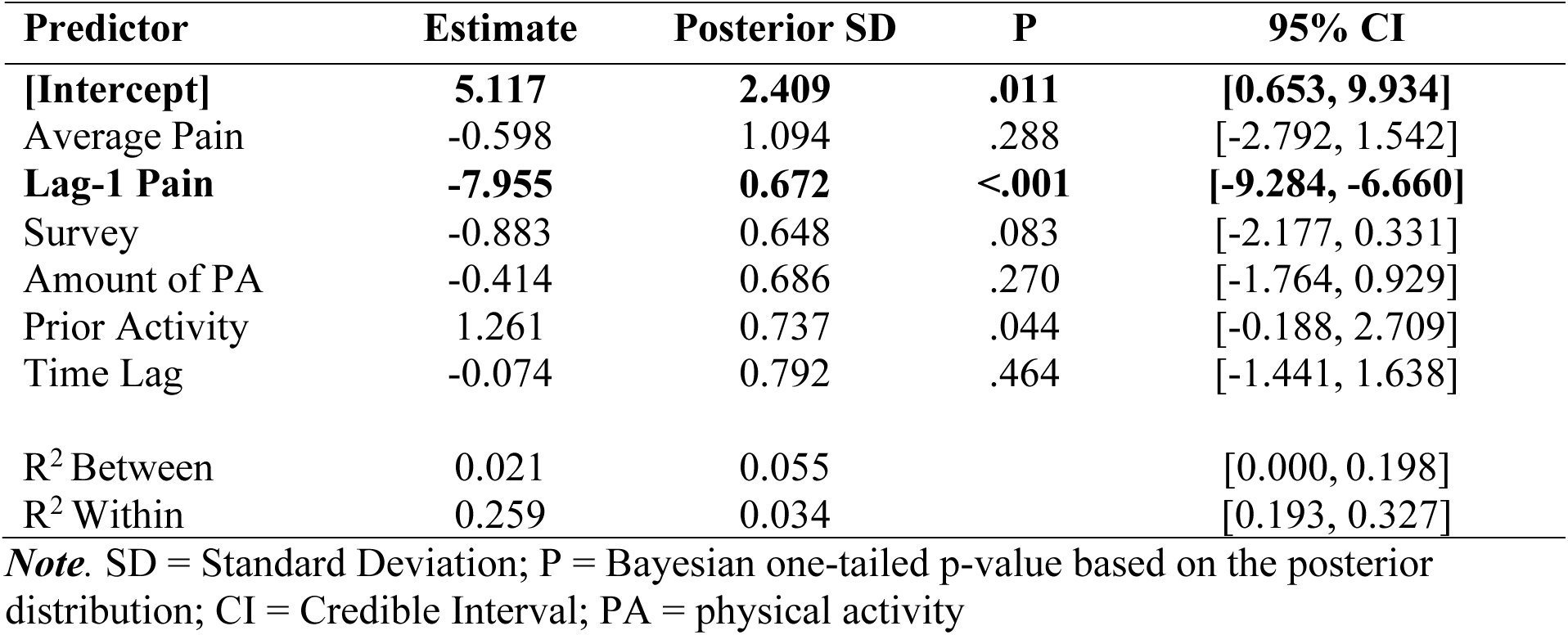
Multivariate predictors of MEP change scores (N = 85, T = 496)

| Predictor | Estimate | Posterior SD | P | 95% CI |
| --- | --- | --- | --- | --- |
| **[Intercept]** | **5.117** | **2.409** | **.011** | **[0.653, 9.934]** |
| Average Pain | -0.598 | 1.094 | .288 | [-2.792, 1.542] |
| **Lag-1 Pain** | **-7.955** | **0.672** | **<.001** | **[-9.284, -6.660]** |
| Survey | -0.883 | 0.648 | .083 | [-2.177, 0.331] |
| Amount of PA | -0.414 | 0.686 | .270 | [-1.764, 0.929] |
| Prior Activity | 1.261 | 0.737 | .044 | [-0.188, 2.709] |
| Time Lag | -0.074 | 0.792 | .464 | [-1.441, 1.638] |
| R<sup>2</sup> Between | 0.021 | 0.055 |  | [0.000, 0.198] |
| R<sup>2</sup> Within | 0.259 | 0.034 |  | [0.193, 0.327] |

**Note.** SD = Standard Deviation; P = Bayesian one-tailed p-value based on the posterior distribution; CI = Credible Interval; PA = physical activity

### 3.8. Variability in PAR

EMA pain observations were marked as indexing PAR if they were not preceded a 6+ minute activity bout, based on the step count threshold. Across all PAR observations, most were preceded by <1 average spm (31%) or 1-5 average spm (32%; Figure 5A) in the 30 minutes before EMA. Even when participants were completely sedentary prior to PAR ratings, approximately 21% of observations exceeded 80/100 indicating severe pain (Figure 5B; Table S6). Distributions were similar for lag-1 pain observations marked as indexing PAR in the main analysis (Figure 5C and Figure 5D).

**Figure 5.**
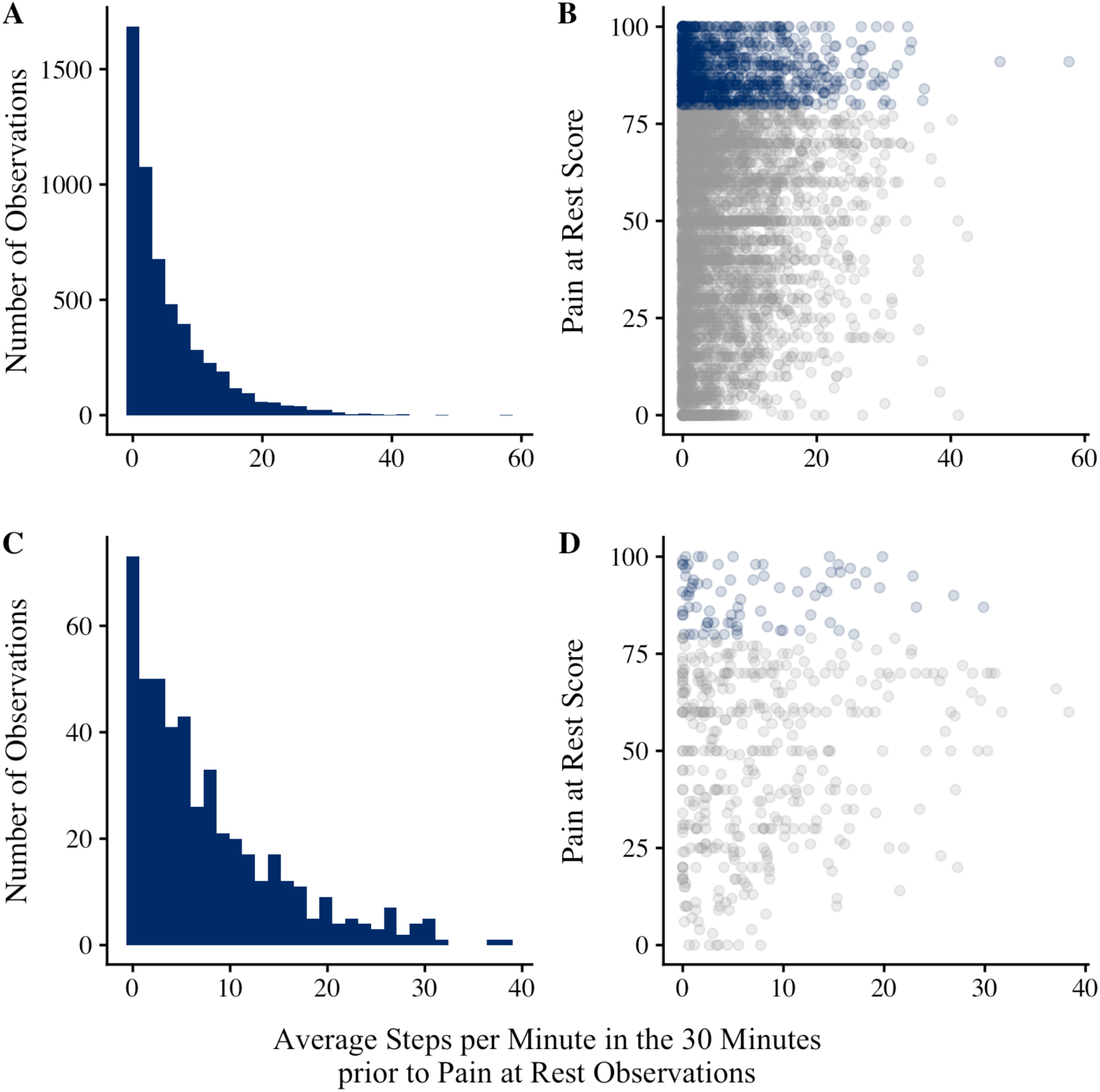
Panel **A** is a histogram describing step counts prior to all EMA pain observations marked as indexing pain at rest (PAR). EMA pain observations were marked as indexing PAR if they were not preceded a 6+ minute activity bout, based on the step count threshold (here, 70spm). Panel **B** is a scatter plot of step counts (x) and PAR scores (y). Blue dots = severe pain (≥80 out of 100). Panels **C** and **D** pertain to lag-1 pain observations marked as indexing PAR in the main analysis.

## 4. Discussion

This study leveraged digital technology to assess movement-evoked pain (MEP) under free-living conditions among adults with moderate-to-severe chronic pain scheduled to receive elective lumbar/thoracolumbar fusion surgery. Numerous studies have assessed MEP as absolute post-activity pain ratings, such that individuals who report higher post-activity pain are considered to have greater MEP. Absolute post-activity pain scores had good reliability.

However, there was a high degree of overlap between post-activity pain ratings and average pain levels calculated from EMA pain ratings, which reduce risk of recall bias associated with retrospective pain reports^13^. Post-activity pain ratings also appeared elevated when pre-activity pain was elevated relative to the individual’s average. Post-activity pain ratings therefore appear to capture overall disability and day-to-day fluctuations in pain, rather than the relationship between movement and pain.

Change scores are commonly used to isolate MEP from PAR by subtracting PAR from post-activity pain ratings. In the current study, MEP change scores had poor reliability when assessed naturalistically and over time. MEP change scores were highly influenced by lag-1 pain observations meant to mimic PAR, such that change scores were lower when lag-1 pain observations were elevated. This is suggestive of ceiling effects, wherein individuals experience lower MEP change scores because PAR is high. Some lag-1 pain observations may not truly capture PAR, as participants were not instructed to rest prior to the EMA pain rating. However, under these naturalistic conditions, wide variability in PAR reports was observed, including when no or very little activity was detected in the 30 minutes prior to PAR observations.

Findings suggest ceiling effects are common, especially when demand characteristics are absent, as over 20% of pain reports following true resting periods indicated severe pain.

Relatively little research has investigated temporal aspects of MEP, including whether MEP is distinct from delayed-onset soreness that may occur hours to days after physical activity^2^.

Findings of the current study suggest MEP may not be temporally bound within a short window after pain-evoking movement. Post-activity pain ratings and MEP change scores collected immediately after or during activity bouts appear largely similar to ratings collected up to 3 hours after activity. The only exceptions were observed at lower step count thresholds (e.g., 50-60spm), and effects were not substantiated in multivariate models. Future studies can utilize event-contingent monitoring. Traditionally, event-contingent monitoring has relied on participants to report that an event (i.e., physical activity) has occurred, in order to receive follow-up prompts^50^. Participants may forget or be unmotivated to complete event-contingent reports, leading to lower compliance and unrepresentative data. Instead, sensor data can be integrated into the EMA platform to automatically trigger prompts after bouts of physical activity^51^. Repeated prompts following bouts of physical activity would be useful for characterizing the rate of decay in pain reports. Additionally, because only examined fixed effects were examined, it remains possible that the temporality of MEP (e.g., rate of decay) varies across individuals, which may have meaningful implications for phenotyping and treatment personalization.

This study has several strengths. Variability in two common operationalizations of MEP across days and times of day were assessed among individuals with moderate-to-severe chronic pain. Additionally, factors that may influence both absolute post-activity pain and MEP change scores when assessed only once were identified, filling known gaps in the growing literature on MEP^2,3^. Participants were largely adherent to the assessment procedures, completing over 84% of EMAs on average and wearing the Fitbit for several weeks. A novel framework to evaluate MEP ecologically by extracting pain ratings following physical activity bouts lasting at least 6 minutes was developed and deployed. By mimicking the 6MWT under free-living conditions, demand characteristics were attenuated, as participants were not aware that their Fitbit and EMA data would be combined to evaluate MEP.

This study also has limitations. The sample was predominantly White and middle-aged. In studies of knee osteoarthritis and nonspecific chronic low back pain, individuals who identified as Black/African American tend to experience greater movement-evoked pain than non-Hispanic White adults^52,53^. Furthermore, movement-evoked pain was more strongly associated with physical and psychosocial functioning among Black/African American adults in these samples^52–55^. Spine surgery is most common among middle-aged and older adults, and over 80% of patients identify as White and non-Hispanic^56^. Thus, larger samples are needed to examine potential interactions between race and movement-evoked pain, including potential variation in predictors of movement-evoked pain and differential associations with surgical outcomes. Face-to-face recruitment may increase sample diversity^57^.

Consumer-grade wearable devices were used to increase feasibility. Fitbits are the most commonly used wearable devices in research settings^28^. Although several studies find that Fitbits accurately measure steps, there is concern about overestimation under free-living conditions^28,38,58^. At present, moderate-intensity physical activity is not well-defined among adults with moderate-to-severe pain interference and disability. Several step count thresholds ranging from low-intensity activity bouts (e.g., 50 steps per minute) to the recommend 100 steps per minute benchmark for moderate intensity exercise were considered^40^. Less than half the sample had any 6-minute activity bouts at the 100 steps per minute threshold. Analysis of heart rate data suggests step count thresholds well below 100 steps per minute are sufficient for indexing moderate levels of physical activity. Importantly, analyses also suggested that factors including time of day and additive effects of activity may impact MEP outcomes. However, these results were not robust across step count thresholds. Further research is needed to define moderate-intensity exercise in this population.

Given observed variability in MEP, especially MEP change scores, continued development of digital methodologies for assessing MEP is recommended. Prior studies suggest that the within-person relationship between physical activity and pain is complex, with a number of factors including overall activity levels, genotype, and psychosocial factors moderating relationships across persons^59–61^. Thus, further within-person assessment is recommended, especially using methods that consider how relationships between physical activity and pain vary across persons^61^. In the spine surgery literature, there is scant investigation of movement-evoked pain, despite *performance* on related tasks (e.g., six-minute walk, timed up and go) appearing as predictors of surgical outcomes^62,63^. Though it is assumed that pain is a primary driver of physical performance, direct investigation of this hypothesis is warranted.

To this end, a dynamic index of MEP based on continuous Fitbit and EMA data was previously developed by the research team^7^. Using multilevel dynamic structural equation modeling^64^, estimates of the strength of association between physical activity and pain was extracted for each individual. This novel framework disentangles MEP from average pain, as someone with low average pain can still experience reliable increases in pain following activity. Additionally, the approach utilizes all available data, instead of only occasions when the individual completes a standardized task or engages in activity above a pre-specified threshold. All individuals who provide activity and EMA data have an MEP estimate indexing the degree to which pain increases with activity, after accounting for important covariates (e.g., time of day). In preliminary analyses, this dynamic index of MEP was a predictor of one-month spine surgery outcomes^7^.

Digital tools can also be used to administer task-based assessment of MEP repeatedly and under naturalistic settings. A smartphone app version of the 6MWT is increasingly being used to evaluate functional impairment^65–68^. Future studies can use this approach to assess MEP by collecting pain ratings before and after the remotely delivered 6MWT. This procedure can be repeated across multiple days and at different times per day to further improve understanding of fluctuations in MEP. Additionally, remote protocols can be developed to capture pain in response to physical activities outside of walking. Although the 6MWT is a common MEP task, other movements (e.g., chair rises, trunk rotation, and standing forward reach) are also relevant to MEP in back pain patients^4^. Some more recent studies define MEP as the aggregate of post-activity pain ratings across several tasks, minus the pre-activity pain rating^17,69^. This procedure takes into account resting pain but will be less subject to ceiling effects given that post-activity pain ratings are weighted more heavily than pre-activity pain.

Overall, this study highlights the need for further research to evaluate the reliability and construct validity of commonly used methods for assessing MEP. The two approaches described above are likely most effective when used together to capture individual differences in the dynamic relationship between activity and pain, and to compare these patterns with task-based MEP assessments delivered remotely and repeated across multiple days and times of day.

Phenotyping chronic pain remains a significant challenge, but improving the precision of MEP measurement may enhance efforts to identify meaningful subgroups within heterogeneous chronic pain populations.

## Data Availability

All data produced in the present study are available upon reasonable request to the authors

## Author Contributions

MF: Conceptualization, Methodology, Formal analysis, Writing – original draft. JZ: Data curation, Investigation, Writing – review & editing.

ZX: Data curation, Investigation, Writing – review & editing. SY: Data curation, Investigation, Writing – review & editing. BB: Data curation, Investigation, Writing – review & editing. SJ: Data curation, Investigation, Writing – review & editing. JZ: Data curation, Investigation, Writing – review & editing. KB: Data curation, Investigation, Writing – review & editing. BG: Supervision, Writing – review & editing. CL: Supervision, Funding acquisition, Writing – review & editing. WR: Supervision, Funding acquisition, Writing – review & editing. JG: Supervision, Funding acquisition, Writing – review & editing.

## Footnotes

1 There were 2 individuals for whom errors in EMA set up led to surveys being administered less than 3 hours apart. Surveys received less than 3 hours after the prior survey were removed from the data set so that lag-1 EMA consistently corresponded to surveys completed approximately 3 hours prior.

## References

1. Breivik H, Borchgrevink PC, Allen SM, et al. Assessment of pain. British Journal of Anaesthesia. 2008;101(1):17–24. doi:10.1093/bja/aen103

2. Butera KA, Chimenti RL, Alsouhibani AM, et al. Through the Lens of Movement-Evoked Pain: A Theoretical Framework of the “Pain-Movement Interface” to Guide Research and Clinical Care for Musculoskeletal Pain Conditions. The Journal of Pain. 2024;25(7):104486. doi:10.1016/j.jpain.2024.01.351

3. Corbett DB, Simon CB, Manini TM, George SZ, Riley JL, Fillingim RB. Movement-evoked pain: Transforming the way we understand and measure pain. PAIN. 2019;160(4):757. doi:10.1097/J.PAIN.0000000000001431

4. Fullwood D, Means S, Merriwether EN, Chimenti RL, Ahluwalia S, Booker SQ. Toward understanding movement-evoked pain (MEP) and its measurement: A scoping review. Clin J Pain. 2021;37(1):61–78. doi:10.1097/AJP.0000000000000891

5. Gilron I, Lao N, Carley M, et al. Movement-evoked pain versus pain at rest in postsurgical clinical trials and in meta-analyses: an updated systematic review. Anesthesiology. 2024;140(3):442–449. doi:10.1097/ALN.0000000000004850

6. Srikandarajah S, Gilron I. Systematic review of movement-evoked pain versus pain at rest in postsurgical clinical trials and meta-analyses: A fundamental distinction requiring standardized measurement. PAIN. 2011;152(8):1734–1739. doi:10.1016/j.pain.2011.02.008

7. Greenberg JK, Frumkin M, Xu Z, et al. Preoperative mobile health data improve predictions of recovery from lumbar spine surgery. Neurosurgery. 2024;95(3):617. doi:10.1227/neu.0000000000002911

8. Knox PJ, Simon CB, Pohlig RT, et al. Movement-Evoked Pain Versus Widespread Pain: A Longitudinal Comparison in Older Adults With Chronic Low Back Pain From the Delaware Spine Studies. The Journal of Pain. 2023;24(6):980–990. doi:10.1016/j.jpain.2023.01.012

9. Lozano-Meca JA, Gacto-Sánchez M, Montilla-Herrador J. Movement-evoked pain is not associated with pain at rest or physical function in knee osteoarthritis. European Journal of Pain. 2024;28(6):987–996. doi:10.1002/ejp.2236

10. Sayers A, Wylde V, Lenguerrand E, et al. Rest Pain and Movement-Evoked Pain as Unique Constructs in Hip and Knee Replacements. Arthritis Care & Research. 2016;68(2):237–245. doi:10.1002/acr.22656

11. Riddle DL, Dumenci L. Preoperative measures of pain at rest and movement-evoked pain in knee arthroplasty: Associations with pain and function outcome trajectories from a prospective multicentre longitudinal cohort study. European Journal of Pain. 29(2):e4723. doi:10.1002/ejp.4723

12. Schneider S, Stone AA, Schwartz JE, Broderick JE. Peak and end effects in patients daily recall of pain and fatigue: A within-subjects analysis. Journal of Pain. 2011;12(2):228–235. doi:10.1016/J.JPAIN.2010.07.001

13. Stone AA, Schwartz JE, Broderick JE, Shiffman SS. Variability of momentary pain predicts recall of weekly pain: A consequence of the peak (or salience) memory heuristic. Personality and Social Psychology Bulletin. 2005;31(10):1340–1346. doi:10.1177/0146167205275615

14. Stone AA, Shiffman S. Ecological momentary assessment (EMA) in behavioral medicine. Ann Behav Med. 1994;16(3):199–202. doi:10.1093/abm/16.3.199

15. Crow JA, Joseph V, Miao G, et al. A domain-oriented approach to characterizing movement-evoked pain. PAIN Reports. 2024;9:e1158. doi:10.1097/PR9.0000000000001158

16. Knox PJ, Simon CB, Pohlig RT, et al. A Standardized Assessment of Movement-evoked Pain Ratings Is Associated With Functional Outcomes in Older Adults With Chronic Low Back Pain. The Clinical Journal of Pain. 2022;38(4):241. doi:10.1097/AJP.0000000000001016

17. Simon CB, Hicks GE, Pieper CF, Byers Kraus V, Keefe FJ, Colón-Emeric C. A Novel Movement-Evoked Pain Provocation Test for Older Adults With Persistent Low Back Pain: Safety, Feasibility, and Associations With Self-reported Physical Function and Usual Gait Speed. The Clinical Journal of Pain. 2023;39(4):166. doi:10.1097/AJP.0000000000001101

18. Knox PJ, Simon CB, Pohlig RT, et al. Construct validity of movement-evoked pain operational definitions in older adults with chronic low back pain. Pain Medicine. 2023;24(8):985–992. doi:10.1093/pm/pnad034

19. Frumkin MR, Rodebaugh TL. The role of affect in chronic pain: A systematic review of within-person symptom dynamics. J Psychosom Res. 2021;147:110527. doi:10.1016/J.JPSYCHORES.2021.110527

20. Quartana PJ, Campbell CM, Edwards RR. Pain catastrophizing: A critical review. Expert Review of Neurotherapeutics. 2009;9(5):745–758. doi:10.1586/ern.09.34

21. Goossens Z, Van Stallen A, Vermuyten J, et al. Day-to-day associations between pain intensity and sleep outcomes in an adult chronic musculoskeletal pain population: A systematic review. Sleep Medicine Reviews. 2025;79:102013. doi:10.1016/j.smrv.2024.102013

22. Knezevic NN, Nader A, Pirvulescu I, Pynadath A, Rahavard BB, Candido KD. Circadian pain patterns in human pain conditions – A systematic review. Pain Practice. 2023;23(1):94–109. doi:10.1111/papr.13149

23. Mun CJ, Burgess HJ, Sears DD, et al. Circadian Rhythm and Pain: a Review of Current Research and Future Implications. Curr Sleep Medicine Rep. 2022;8(4):114–123. doi:10.1007/s40675-022-00228-3

24. Lambin DI, Thibault P, Simmonds M, Lariviere C, Sullivan MJL. Repetition-induced activity-related summation of pain in patients with fibromyalgia. PAIN. 2011;152(6):1424–1430. doi:10.1016/j.pain.2011.02.030

25. Sullivan MJL, Larivière C, Simmonds M. Activity-related summation of pain and functional disability in patients with whiplash injuries. PAIN®. 2010;151(2):440–446. doi:10.1016/j.pain.2010.08.005

26. Sullivan MJL, Thibault P, Andrikonyte J, Butler H, Catchlove R, Larivière C. Psychological influences on repetition-induced summation of activity-related pain in patients with chronic low back pain. PAIN®. 2009;141(1):70–78. doi:10.1016/j.pain.2008.10.017

27. Greenberg JK, Frumkin MR, Javeed S, et al. Feasibility and acceptability of a preoperative multimodal mobile health assessment in spine surgery candidates. Neurosurgery. 2023;92(3):538–546. doi:10.1227/neu.0000000000002245

28. Fuller D, Colwell E, Low J, et al. Reliability and validity of commercially available wearable devices for measuring steps, energy expenditure, and heart rate: Systematic review. JMIR mHealth and uHealth. 2020;8(9):e18694. doi:10.2196/18694

29. Ader DN. Developing the Patient-Reported Outcomes Measurement Information System (PROMIS). Medical Care. 2007;45(5):S1. doi:10.1097/01.mlr.0000260537.45076.74

30. Amtmann D, Cook KF, Jensen MP, et al. Development of a PROMIS item bank to measure pain interference. PAIN. 2010;150(1):173–182. doi:10.1016/j.pain.2010.04.025

31. Stone AA, Broderick JE, Junghaenel DU, Schneider S, Schwartz JE. PROMIS fatigue, pain intensity, pain interference, pain behavior, physical function, depression, anxiety, and anger scales demonstrate ecological validity. Journal of Clinical Epidemiology. 2016;74:194–206. doi:10.1016/J.JCLINEPI.2015.08.029

32. Yates T, Henson J, McBride P, et al. Moderate-intensity stepping in older adults: insights from treadmill walking and daily living. International Journal of Behavioral Nutrition and Physical Activity. 2023;20(1):31. doi:10.1186/s12966-023-01429-x

33. Frumkin MR, Greenberg JK, Boyd P, et al. Establishing the reliability, validity, and prognostic utility of the momentary pain catastrophizing scale for use in ecological momentary assessment research. J Pain. 2023;28(8):1423–1433. doi:10.1016/j.jpain.2023.03.010

34. Mehl MR, Conner TS. Handbook of Research Methods for Studying Daily Life. Guilford Publications; 2013.

35. Palada V, Gilron I, Canlon B, Svensson CI, Kalso E. The circadian clock at the intercept of sleep and pain. PAIN. 2020;161(5):894. doi:10.1097/j.pain.0000000000001786

36. Ham SA, Reis JP, Strath SJ, Dubose KD, Ainsworth BE. Discrepancies between methods of identifying objectively determined physical activity. Med Sci Sports Exerc. 2007;39(1):52–58. doi:10.1249/01.mss.0000235886.17229.420

37. Silva GS, Yang H, Collins JE, Losina E. Validating Fitbit for Evaluation of Physical Activity in Patients with Knee Osteoarthritis: Do Thresholds Matter? ACR Open Rheumatol. 2019;1(9):585–592. doi:10.1002/acr2.11080

38. Redenius N, Kim Y, Byun W. Concurrent validity of the Fitbit for assessing sedentary behavior and moderate-to-vigorous physical activity. BMC Medical Research Methodology. 2019;19(1):29. doi:10.1186/s12874-019-0668-1

39. Roberts-Lewis SF, White CM, Ashworth M, Rose MR. Validity of Fitbit activity monitoring for adults with progressive muscle diseases. Disability and Rehabilitation. 2022;44(24):7543–7553. doi:10.1080/09638288.2021.1995057

40. Tudor-Locke C, Craig CL, Aoyagi Y, et al. How many steps/day are enough? For older adults and special populations. Int J Behav Nutr Phys Act. 2011;8(1):80. doi:10.1186/1479-5868-8-80

41. Barnett A, van den Hoek D, Barnett D, Cerin E. Measuring moderate-intensity walking in older adults using the ActiGraph accelerometer. BMC Geriatr. 2016;16(1):211. doi:10.1186/s12877-016-0380-5

42. Hall KS, Howe CA, Rana SR, Martin CL, Morey MC. METs and Accelerometry of Walking in Older Adults: Standard versus Measured Energy Cost. Med Sci Sports Exerc. 2013;45(3):574–582. doi:10.1249/MSS.0b013e318276c73c

43. Tanaka H, Monahan KD, Seals DR. Age-predicted maximal heart rate revisited. J Am Coll Cardiol. 2001;37(1):153–156. doi:10.1016/s0735-1097(00)01054-8

44. Medicine AC of S. ACSM’s Guidelines for Exercise Testing and Prescription. Lippincott Williams & Wilkins; 2014.

45. Kuznetsova A, Brockhoff PB, Christensen RH. lmerTest package: tests in linear mixed effects models. Journal of statistical software. 2017;82:1–26.

46. Slipetz LR, Falk A, and Henry TR. Missing Data in Discrete Time State-Space Modeling of Ecological Momentary Assessment Data: A Monte-Carlo Study of Imputation Methods. Multivariate Behavioral Research. 2025;60(4):695–710. doi:10.1080/00273171.2025.2469055

47. Daniels MJ, Hogan JW. Missing Data in Longitudinal Studies: Strategies for Bayesian Modeling and Sensitivity Analysis. Chapman and Hall/CRC; 2008. doi:10.1201/9781420011180

48. McNeish D, Hamaker EL. A Primer on Two-Level Dynamic Structural Equation Models for Intensive Longitudinal Data in Mplus. Psychological Methods. Published online 2019. doi:10.1037/MET0000250

49. Boonstra AM, Stewart RE, Köke AJA, et al. Cut-Off Points for Mild, Moderate, and Severe Pain on the Numeric Rating Scale for Pain in Patients with Chronic Musculoskeletal Pain: Variability and Influence of Sex and Catastrophizing. Front Psychol. 2016;7:1466. doi:10.3389/fpsyg.2016.01466

50. Stone AA, Obbarius A, Junghaenel DU, Wen CKF, Schneider S. High-resolution, field approaches for assessing pain: Ecological Momentary Assessment. Pain. 2021;162(1):4–9. doi:10.1097/j.pain.0000000000002049

51. Dunton GF, Dzubur E, Intille S. Feasibility and Performance Test of a Real-Time Sensor-Informed Context-Sensitive Ecological Momentary Assessment to Capture Physical Activity. Journal of Medical Internet Research. 2016;18(6):e5398. doi:10.2196/jmir.5398

52. Booker S, Cardoso J, Cruz-Almeida Y, et al. Movement-evoked pain, physical function, and perceived stress: An observational study of ethnic/racial differences in aging non-Hispanic Blacks and non-Hispanic Whites with knee osteoarthritis. Experimental Gerontology. 2019;124:110622. doi:10.1016/j.exger.2019.05.011

53. Penn TM, Overstreet DS, Aroke EN, et al. Perceived injustice helps explain the association between chronic pain stigma and movement-evoked pain in adults with nonspecific chronic low back pain. Pain medicine. 2020;21(11):3161–3171.

54. Bartley EJ, Hossain NI, Gravlee CC, et al. Race/Ethnicity Moderates the Association Between Psychosocial Resilience and Movement-Evoked Pain in Knee Osteoarthritis. ACR Open Rheumatology. 2019;1(1):16–25. doi:10.1002/acr2.1002

55. Morais C, Palit S, Cardoso J, et al. Movement-Evoked pain is differentially associated with physical and psychological functioning in non-Hispanic Blacks vs. non-Hispanic Whites. The Journal of Pain. 2021;22(5):609–610. doi:10.1016/j.jpain.2021.03.125

56. Al Jammal OM, Shahrestani S, Delavar A, et al. Demographic predictors of treatments and surgical complications of lumbar degenerative diseases: An analysis of over 250,000 patients from the National Inpatient Sample. Medicine. 2022;101(11):e29065. doi:10.1097/MD.0000000000029065

57. Carter CR, Maki J, Ackermann N, Waters EA. Inclusive Recruitment Strategies to Maximize Sociodemographic Diversity among Participants: A St. Louis Case Study. MDM Policy Pract. 2023;8(1):23814683231183646. doi:10.1177/23814683231183646

58. Feehan LM, Geldman J, Sayre EC, et al. Accuracy of Fitbit Devices: Systematic Review and Narrative Syntheses of Quantitative Data. JMIR mHealth and uHealth. 2018;6(8):e10527. doi:10.2196/10527

59. Davis TJ, Hevel DJ, Dunton GF, Maher JP. Bidirectional associations between physical activity and pain among older adults: an ecological momentary assessment study. Journal of aging and physical activity. 2022;31(2):240–248.

60. Martire LM, Wilson SJ, Small BJ, Conley YP, Janicki PK, Sliwinski MJ. COMT and OPRM1 genotype associations with daily knee pain variability and activity induced pain. Scandinavian Journal of Pain. 2016;10(1):6–12. doi:10.1016/j.sjpain.2015.07.004

61. Tynan M, Virzi N, Wooldridge JS, Morse JL, Herbert MS. Examining the Association Between Objective Physical Activity and Momentary Pain: A Systematic Review of Studies Using Ambulatory Assessment. The Journal of Pain. 2024;25(4):862–874. doi:10.1016/j.jpain.2023.10.021

62. Komodikis G, Gannamani V, Neppala S, Li M, Merli GJ, Harrop JS. Usefulness of Timed Up and Go (TUG) test for prediction of adverse outcomes in patients undergoing thoracolumbar spine surgery. Neurosurgery. 2020;86(3):E273–E280.

63. Takenaka H, Kamiya M, Sugiura H, Nishihama K, Suzuki J, Hanamura S. Minimal Clinically Important Difference of the 6-Minute Walk Distance in Patients Undergoing Lumbar Spinal Canal Stenosis Surgery: 12 Months Follow-Up. Spine. 2023;48(8):559. doi:10.1097/BRS.0000000000004566

64. Asparouhov T, Hamaker EL, Muthén B. Dynamic structural equation models. Struct Equ Modeling. 2018;25(3):359–388. doi:10.1080/10705511.2017.1406803

65. Maldaner N, Sosnova M, Zeitlberger AM, et al. Evaluation of the 6-minute walking test as a smartphone app-based self-measurement of objective functional impairment in patients with lumbar degenerative disc disease. Journal of Neurosurgery. 2020;33(6):779–788. doi:10.3171/2020.5.SPINE20547

66. Salvi D, Poffley E, Orchard E, Tarassenko L. The Mobile-Based 6-Minute Walk Test: Usability Study and Algorithm Development and Validation. JMIR mHealth and uHealth. 2020;8(1):e13756. doi:10.2196/13756

67. Schubert C, Archer G, Zelis JM, et al. Wearable devices can predict the outcome of standardized 6-minute walk tests in heart disease. npj Digit Med. 2020;3(1):92. doi:10.1038/s41746-020-0299-2

68. Simmich J, Andrews NE, Claus A, Murdoch M, Russell TG. Assessing a GPS-Based 6-Minute Walk Test for People With Persistent Pain: Validation Study. JMIR Formative Research. 2024;8(1):e46820. doi:10.2196/46820

69. Kahan R, Woznowski-Vu A, Huebner JL, et al. Psychological and immunological associations with movement-evoked low back pain among older adults. PAIN Reports. 2025;10(3):e1262. doi:10.1097/PR9.0000000000001262

