## Supplementary materials for "Rethinking Measurement of Movement-Evoked Pain with Digital Technology"

**Fig S1**. Impact of 6-minute step count thresholds on heart rate verification and participants included

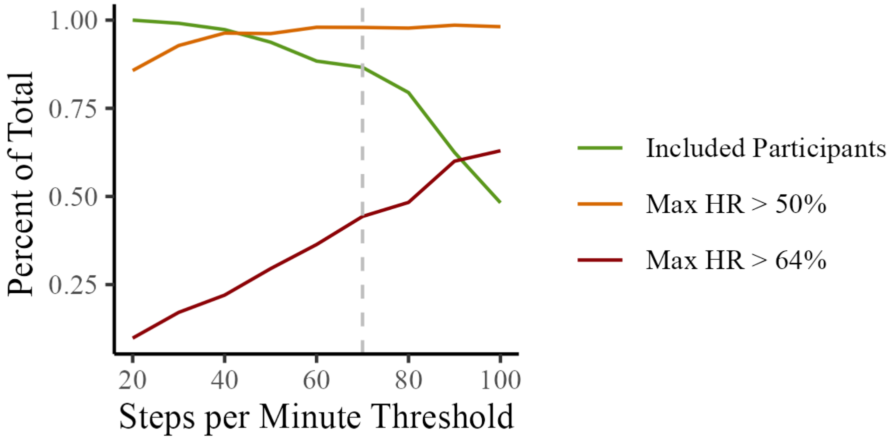


*Note*. A threshold of 70 steps per minute (grey dotted line) was retained as this threshold minimized participant exclusion (green line) due to a lack of observations, while maximizing heart rate (HR) verification of exercise intensity at 50% of max HR (orange line) and 64% of max HR (red line) thresholds.

**Table S1**. Reliability of MEP across step count thresholds

| Steps per Minute Threshold | Absolute Post-Activity Pain | | MEP Change Scores | |
|---|---|---|---|---|
| | M (SD) of Within-Person Variability | ICC | M (SD) of Within-Person Variability | ICC |
| 50 | 12.23 (8.62) | 0.72 | 13.64 (11.38) | 0.04 |
| 60 | 11.6 (9.33) | 0.74 | 13.35 (10.67) | 0.05 |
| 70 | 10.14 (6.64) | 0.76 | 11.58 (7.78) | 0.08 |
| 80 | 10.76 (6.84) | 0.76 | 12.49 (8.16) | 0.06 |
| 90 | 10.66 (7.41) | 0.74 | 11.58 (8.35) | 0.07 |
| 100 | 12.5 (8.49) | 0.68 | 13.23 (9.15) | 0.11 |

*Note*. M = mean; SD = standard deviation; ICC = intraclass correlation coefficient

**Table S2**. Univariate predictors of post-activity pain ratings across step count thresholds

| Steps per Minute Threshold | Average Pain | Lag-1 Pain | Survey | Survey$^2$ | Amount of PA | Prior Activity | Time Lag |
|---|---|---|---|---|---|---|---|
| 50 | 22.07 (0.48)* | 3.58 (0.49)* | 0.93 (0.33)* | -0.91 (0.28)* | 0.14 (0.42) | 0.30 (0.43) | -0.14 (0.52) |
| 60 | 22.53 (0.55)* | 2.89 (0.56)* | 0.40 (0.38) | -0.87 (0.31)* | 0.21 (0.47) | 0.52 (0.48) | -0.61 (0.57) |
| 70 | 22.46 (0.72)* | 3.00 (0.62)* | 0.43 (0.42) | -0.64 (0.35) | -0.04 (0.51) | 0.45 (0.53) | -0.08 (0.62) |
| 80 | 22.70 (0.79)* | 2.47 (0.73)* | 0.33 (0.50) | -0.90 (0.41)* | -0.23 (0.60) | 0.58 (0.61) | -0.87 (0.75) |
| 90 | 22.95 (0.88)* | 1.96 (0.90)* | 0.47 (0.60) | -1.00 (0.49)* | -0.33 (0.74) | 0.37 (0.76) | 1.23 (0.95) |
| 100 | 22.93 (1.28)* | 2.66 (1.37) | 0.48 (0.81) | -0.80 (0.70) | 0.41 (1.05) | 1.36 (1.09) | 0.92 (1.42) |

*Note*. Model results are presented as coefficient (standard error). * = p < .05; PA = physical activity

**Table S3**. Multivariate predictors of post-activity pain ratings across step count thresholds

| Steps per Minute Threshold | Average Pain | Lag-1 Pain | Survey$^2$ | Amount of PA | Prior Activity | Time Lag |
|---|---|---|---|---|---|---|
| 50 | 21.87 (0.50)* | 3.99 (0.51)* | -1.07 (0.26)* | -0.30 (0.46) | 0.54 (0.47) | -0.61 (0.52) |
| 60 | 22.41 (0.59)* | 3.27 (0.58)* | -1.00 (0.30)* | -0.28 (0.51) | 0.86 (0.52) | -0.99 (0.58) |
| 70 | 22.31 (0.76)* | 3.31 (0.64)* | -0.63 (0.33) | -0.56 (0.57) | 1.12 (0.58) | -0.42 (0.62) |
| 80 | 22.43 (0.88)* | 3.02 (0.76)* | -0.87 (0.39)* | -0.73 (0.67) | 1.29 (0.70) | -1.00 (0.76) |
| 90 | 22.73 (0.97)* | 2.24 (0.93)* | -0.76 (0.46) | -0.64 (0.80) | 0.91 (0.80) | 0.64 (0.96) |
| 100 | 22.65 (1.41)* | 3.05 (1.30)* | -0.80 (0.62) | -0.63 (1.15) | 1.71 (1.17) | 0.50 (1.43) |

*Note*. Model results are presented as coefficient (posterior standard deviation). * = 95% credible interval does not contain zero; PA = physical activity

**Table S4**. Univariate predictors of MEP change scores across step count thresholds

| Steps per Minute Threshold | Average Pain | Lag-1 Pain | Survey | Survey$^2$ | Amount of PA | Prior Activity | Time Lag |
|---|---|---|---|---|---|---|---|
| 50 | -0.20 (0.72) | -7.78 (0.53)* | -1.03 (0.71) | -0.80 (0.60) | 0.59 (0.61) | 1.25 (0.62)* | -2.44 (0.70)* |
| 60 | 0.09 (0.88) | -8.64 (0.61)* | -1.79 (0.84)* | -0.55 (0.70) | 0.34 (0.71) | 2.13 (0.72)* | -2.39 (0.82)* |
| 70 | -0.53 (0.99) | -8.17 (0.66)* | -1.82 (0.92)* | 0.23 (0.75) | -0.31 (0.77) | 2.05 (0.78)* | -1.08 (0.86) |
| 80 | -0.07 (1.09) | -8.43 (0.78)* | -1.54 (1.11) | -0.49 (0.89) | -0.32 (0.92) | 1.77 (0.92) | -1.74 (1.03) |
| 90 | 1.34 (1.27) | -8.79 (0.94)* | -0.57 (1.38) | -0.28 (1.04) | -2.16 (1.13) | 0.84 (1.12) | -0.38 (1.24) |
| 100 | 2.25 (1.99) | -9.90 (1.43)* | 1.16 (1.97) | -1.59 (1.51) | -3.36 (1.71) | 0.10 (1.70) | -0.71 (1.80) |

*Note*. Model results are presented as coefficient (standard error). * = p < .05; PA = physical activity

**Table S5**. Multivariate predictors of MEP change scores across step count thresholds

| Steps per Minute Threshold | Average Pain | Lag-1 Pain | Survey | Amount of PA | Prior Activity | Time Lag |
|---|---|---|---|---|---|---|
| 50 | -0.19 (0.82) | -7.55 (0.53)* | -0.46 (0.53) | -0.19 (0.58) | 0.91 (0.62) | -1.59 (0.67)* |
| 60 | 0.13 (0.96) | -8.38 (0.62)* | -1.00 (0.60) | -0.18 (0.64) | 1.43 (0.68)* | -1.00 (0.76) |
| 70 | -0.60 (1.09) | -7.96 (0.67)* | -0.88 (0.65) | -0.41 (0.69) | 1.26 (0.74) | 0.07 (0.79) |
| 80 | -0.17 (1.19) | -8.08 (0.79)* | -1.18 (0.76) | -0.72 (0.83) | 1.11 (0.88) | -0.78 (0.98) |
| 90 | 1.29 (1.41) | -8.51 (0.95)* | -0.30 (0.94) | -1.31 (0.95) | 0.96 (1.03) | 1.32 (1.23) |
| 100 | 2.11 (2.26) | -9.39 (1.46)* | 0.99 (1.39) | -1.46 (1.49) | 0.43 (1.66) | -0.12 (1.81) |

*Note*. Model results are presented as coefficient (posterior standard deviation). * = 95% credible interval does not contain zero; PA = physical activity

**Table S6.** Descriptives of resting pain ratings by pre-EMA activity levels.

| Steps per Minute Threshold | Number of Observations | Percent of Total | Median (SD) Pain Rating | % Severe Pain |
|---|---|---|---|---|
| <1 spm | 1667 | 31% | 56.0 (27.7) | 20.8% |
| 1-5 spm | 1732 | 32% | 59.5 (27.0) | 19.4% |
| 5-10 spm | 1030 | 19% | 60.0 (25.9) | 21.5% |

*Note*. SD = standard deviation; severe pain cut-off = 80/100
